# Examining the Neural and Behavioral Impact of Accelerated Intermittent Theta Burst Stimulation (iTBS) in People with Opioid Use Disorder (OUD) Who Smoke Tobacco Cigarettes: A Pilot Study

**DOI:** 10.1101/2025.09.10.25335485

**Authors:** Dylan H. Ballard, Thomas G. Adams, Rajendra A. Morey, Rebika Khanal, Seth S. Himelhoch, Gopalkumar Rakesh

## Abstract

Novel therapies are needed to improve smoking cessation outcomes in people with opioid use disorder (OUD), as they are far more likely to smoke cigarettes (70-90%) compared to the general population (11.6%) and demonstrate poorer response to smoking cessation interventions. This pilot study was the first to explore the impact of a single day (four sessions) of accelerated intermittent theta burst stimulation (iTBS) (1800 pulses/session) versus sham iTBS on the left dorsolateral prefrontal cortex (L.dlPFC) in people with OUD who smoke tobacco cigarettes (n=8 received iTBS, n=7 received sham iTBS). Resting state functional connectivity (rsFC) was acquired at baseline and after the fourth session. Attentional bias for cigarette and opioid cues, and craving assessments were completed at baseline, and after the first and fourth sessions.

Connectivity between the L.dlPFC seed and a cluster comprising the left anterior supramarginal gyrus (SMG) showed a significant group × time interaction, with planned comparisons showing a greater increase at follow-up with iTBS compared to sham iTBS (*t_12_*=6.37, *beta*=0.40, *p*<0.001). Cigarette cue attentional bias showed a significant group × session interaction (*t_82_*=2.34*, p*=0.02), with planned comparisons revealing a decrease after iTBS and an increase after sham iTBS. No effect of iTBS was observed for opioid cue attentional bias. Cigarette craving decreased with both iTBS and sham iTBS but did not show a significant group × session interaction. These results are promising but need to be interpreted with caution, given the limited sample size and multiple comparisons. Future trials could examine the effects of increased doses of iTBS (e.g., more days of accelerated iTBS) to identify the dosing required to promote smoking cessation among individuals with OUD effectively.

## Introduction

Approximately 12% of adults in the United States smoke tobacco cigarettes (Wang et al., 2018). People with opioid use disorder (OUD) smoke cigarettes at a much higher rate (i.e., 70-90%) (Guydish et al., 2016, Rajabi et al., 2019). People with OUD and who smoke cigarettes report higher opioid craving, are opioid dependent, and are more challenging to retain in treatment compared to those with OUD who do not smoke cigarettes (Lichenstein et al., 2019, Mannelli et al., 2013). They also demonstrate a poorer response to smoking cessation treatments than people who smoke cigarettes but do not have OUD (Nahvi et al., 2021, Nahvi et al., 2014, Vlad et al., 2020). Novel interventions are needed to optimize smoking cessation treatment in OUD (Lichenstein et al., 2019).

Resting state functional connectivity (rsFC) studies have shown aberrations in the default mode network (DMN), frontoparietal network (FPN), salience network (SN), and visual network (VN) in people who smoke relative to healthy controls (S.J. Brooks, 2017). DMN activity correlates with cigarette craving, and FPN is activated by smoking cessation interventions, which also decrease DMN activity (S.J. Brooks, 2017). Interestingly, the SN mediates the switch between the DMN and FPN (Menon and Uddin, 2010).

People who smoke cigarettes or have OUD demonstrate an attentional bias for the respective substance (opioid or cigarette) cues (MacLean et al., 2018, Masiero et al., 2019). Attentional bias quantifies incentive salience (Berridge and Robinson, 2016) and correlates with the severity of multiple substance use disorders (MacLean et al., 2018, Masiero et al., 2019). Attentional bias is also predictive of response to treatment and relapse (MacLean et al., 2018, Rahmani et al., 2024)..

Transcranial magnetic stimulation (TMS) is a non-invasive neuromodulation modality that has shown efficacy in reducing cigarette craving and use (Mehta et al., 2023, Zangen et al., 2021). The FDA-approved deep TMS protocol for smoking cessation (10 Hz) includes 18 sessions spread over 6 weeks, and each session lasts an hour (Zangen et al., 2021). Intermittent theta burst stimulation (iTBS) is a relatively novel TMS paradigm known for its potency and efficiency (delivery time of 3 to 9 minutes, depending on dose) compared to other repetitive TMS paradigms (Suppa et al., 2016). Previous studies have shown iTBS to decrease tobacco craving (n=4 studies) in people who smoke (Dieler et al., 2014, Mikellides et al., 2022, Rakesh et al., 2024, Upton et al., 2023) and opioid craving (n=2 studies) in people with OUD (Gong et al., 2023, Kang et al., 2022).

Four studies have examined the effects of TMS targeting the left dorsolateral prefrontal cortex (L. dlPFC) on rsFC in individuals who smoke cigarettes (Jordan et al., 2023, Li et al., 2017, Petersen et al., 2025, Rakesh et al., 2024). Only one of these delivered iTBS (1800 pulses at 120% resting motor threshold [RMT]) (Rakesh et al., 2024). Findings from this study suggest that iTBS significantly enhanced rsFC between the L.dlPFC and bilateral medial prefrontal cortex, bilateral temporal poles, and pons compared to sham iTBS (Rakesh et al., 2024). Previous studies have examined the effects of L.dlPFC iTBS (600-1800 pulses/session) on rsFC in healthy participants who do not smoke or use tobacco (Alkhasli et al., 2019, Singh et al., 2020, Tang et al., 2019). These studies showed iTBS to affect rsFC between L.dlPFC and other regions (caudate, insula, amygdala), and higher stimulation intensities affected rsFC between these regions (caudate, insula, amygdala) (Alkhasli et al., 2019, Singh et al., 2020, Tang et al., 2019).

Accelerated iTBS, wherein multiple iTBS sessions (1800 pulses/session) are delivered daily, has shortened time to recovery and increased response rates in major depressive disorder (MDD) (Cole et al., 2022). No studies have explored the impact of iTBS (including accelerated protocols) in people with OUD who smoke. The current pilot study compared the effects of accelerated iTBS versus sham iTBS on 1) rsFC from three seed regions [L.dlPFC, right and left insula] to all other brain voxels (primary outcome), 2) acute measures of cue-based attentional bias, and craving scales for cigarettes and opioids (secondary outcomes). We hypothesized that iTBS would affect rsFC between the L.dlPFC and right/left insulae, given its critical role in cigarette smoking (Naqvi et al., 2007). We also predicted a significant decrease in cigarette craving and cigarette cue attentional bias with iTBS compared with sham iTBS. We did not expect a considerable change in opioid craving or attentional bias, given that these participants were stabilized on medications for OUD when they enrolled in the trial. Exploratory analyses concerned global ROI-to-ROI changes in rsFC.

## Method

### Experimental Protocol

The study protocol was approved by the Medical Institutional Review Board (IRB) at the University of Kentucky (IRB#56570) and was registered on clinicaltrials.gov (NCT05049460). Participants with OUD in remission and smoking tobacco cigarettes were recruited from a specialized OUD clinic within the University of Kentucky Medical Center. Participants were eligible for the study if they were 1) between 18 and 60 years of age, 2) right-handed, 3) able to read, understand, and communicate in English to provide informed consent, 4) stabilized on medications for OUD (methadone, buprenorphine-naloxone, or injectable extended-release naltrexone) for at least two weeks, 5) demonstrating cue-based attentional bias for either cigarette cues or opioid cues, 6) actively smoking cigarettes, and 7) willing to abstain from all substance use during the study period. Participants with any of the following were excluded from study participation: traumatic brain injury, history of seizures or seizure disorder, history of or current diagnosis of schizophrenia, intracranial metal shrapnel, previous adverse effects with TMS, or a positive pregnancy test.

After completing a phone screen to determine eligibility, participants were scheduled for Day 1 procedures, which consisted of informed consent, collection of demographic details, measuring carbon monoxide (CO) using a Smokerlyzer Breath CO Monitor (Bedfont Scientific Ltd., Rochester, England), measuring blood alcohol level using a Breathalyzer, administering the Fagerstrom Test for Nicotine Dependence (FTND) (Heatherton et al., 1991), cue-based attentional bias screen for cigarette and opioid cues, structural MRI, and a functional MRI (fMRI) scan to measure rsFC.

Participants were randomized and scheduled to receive iTBS or sham iTBS a week after Day 1 (Day 2). On Day 2, participants received four sessions of iTBS or sham iTBS at 120% RMT (Rakesh et al., 2024). Sessions were delivered at intervals of 50 minutes, which is consistent with the accelerated protocol for MDD (Cole et al., 2022). The following assessments were administered in the indicated order before iTBS/sham iTBS (baseline) and after the first and the fourth sessions of iTBS/sham iTBS: 1) cigarette cue attentional bias (Rakesh et al., 2023), 2) opioid cue attentional bias, 3) the short form version of tobacco craving questionnaire (TCQ-SF) (Heishman et al., 2008), and 4) the opioid craving scale (OCS) (McHugh et al., 2014). An fMRI scan to measure rsFC was acquired for all participants within 30 minutes of the fourth session of iTBS/sham iTBS.

### Structural and functional MRI

Neuroimaging acquisition was completed with a 3T Siemens Magnetom PRISMA Scanner. Structural MRI images (MPRAGE) were acquired before iTBS. Multi-slice gradient echoplanar imaging (EPI) resting state images were acquired before iTBS/sham iTBS, and within 30 minutes after iTBS/sham iTBS. Details of the structural and fMRI scan sequences are provided in the Supplementary Material Section 1.

### Neuronavigated iTBS

Structural brain MRI images acquired on Day 1 were input into Brainsight software (Rogue Solutions, Montreal, Canada). Consensus Montreal Neurological Institute (MNI) coordinates for Brodmann area 46 were used to identify the L.dlPFC (-44, 40, 29) (Fox et al., 2012, Rajkowska and Goldman-Rakic, 1995, Rakesh et al., 2024). Four sessions of iTBS (1,800 pulses/session) were delivered to this target with a figure-8 coil (MagVenture x100 TMS device, Denmark) at 120% RMT, at intervals of 50 minutes.

### Attentional Bias

Measuring attentional bias with eye tracking can improve reliability and consistency relative to older measurement methods, such as reaction time (Ataya et al., 2012, Field and Christiansen, 2012, Rahmani et al., 2024). Gaze fixation time (in milliseconds) was measured with a Tobii Pro Fusion 120 Hz eye tracker (Rakesh et al., 2023). Participants were exposed to two separate sets of cues – 1) 40 pairs of cigarette cues, consisting of 20 pairs of images of people smoking cigarettes and corresponding neutral cues, and 20 pairs comprised of images of cigarette paraphernalia (e.g., a pack of cigarettes) and corresponding neutral cues, and 2) 40 pairs of opioid cues, consisting of 20 pairs of images of people using opioids and corresponding neutral cues, and 20 pairs of images of opioid paraphernalia (e.g., a needle) and corresponding neutral cues. Each set of cues was mixed with 40 pairs of neutral filler cues. All cue pairs were presented for 2 seconds and separated by a fixation cross of 1 second (Rakesh et al., 2023). Cues were selected from validated databases (Ekhtiari et al., 2020, Manoliu et al., 2021, Rakesh et al., 2023).

### Data analyses

#### Seed-based rsFC analyses

CONN functional connectivity toolbox (version 21a) was used for all preprocessing and neuroimaging analyses (Nieto-Castanon, 2020). Details on preprocessing and denoising are listed in the Supplementary Material Section 2. We used the default parcellation atlas in CONN. For first-level analyses, participant-level data of matrices assessing functional connectivity of three seed regions (L.dlPFC, right insula, and left insula) with all other voxels were estimated. A seed region was constructed for L.dlPFC in CONN using the MNI coordinate –44,29,40 and a sphere radius of 5 mm. Default labels in CONN’s parcellation atlas were chosen as seeds for the right and left insulae seeds.

Functional connectivity strength was represented by Fisher-transformed bivariate correlation coefficients from a weighted general linear model (GLM) (Nieto-Castanon, 2020). These were calculated separately between the three seed regions and every target voxel, characterizing the association between their BOLD signal time series. Individual scans were weighted by a boxcar signal characterizing each time point, convolved with an SPM canonical hemodynamic response function, and rectified.

Second-level analyses were performed using GLM (Worsley et al., 1996). For each voxel, a separate GLM was estimated, with first-level connectivity measures at this connection as dependent variables and independent variables being treatment group (iTBS, sham iTBS), time (before iTBS/sham iTBS, after iTBS/sham iTBS), and the treatment group × time interactions. Voxel-level hypotheses were evaluated using multivariate parametric statistics with random effects across participants and sample covariance estimation across multiple measurements. Inferences were performed at the level of individual clusters (groups of similar connections). Cluster-level inferences were based on parametric statistics from Gaussian Random Field Theory (Worsley et al., 1996) (Nieto-Castanon, 2020). Results were thresholded using a combination of a p < 0.001 voxel-level threshold and a False Discovery Rate (FDR) corrected p value < 0.05 cluster-level threshold (Chumbley et al., 2010).

#### Attentional Bias and Craving

Gaze fixation times were averaged for the two sets of cues across 3-time points (baseline, after the first session of iTBS/sham iTBS, and after four sessions of iTBS/sham iTBS). Cue attentional bias was calculated separately for cigarette and opioid cues by subtracting the fixation time on neutral cues from the fixation time on respective substance cues (Rakesh et al., 2023).

We performed regression analyses using linear mixed effects models (LMEM) to examine group differences in attentional bias and craving changes across the three time points. The LMEMs had the following dependent variables (DVs): 1) cigarette cue attentional bias, 2) opioid cue attentional bias, 3) cigarette craving, and 4) opioid craving. Fixed effects for both models included the treatment group (iTBS vs. sham iTBS), session (baseline, after one session, and after four sessions), and the treatment group × session interaction. The participant identification number was modeled as a random effect for all models. All analyses were completed with MATLAB version R2022a.

#### Correlation between rsFC and behavioral measures

GLM analyses explored the following associations: 1) between changes in seed-based rsFC (three seed regions [the L.dlPFC, right and left insulae] and all voxels) across time, and changes in cue attentional bias for cigarettes and opioids, and 2) between changes in seed-based rsFC across time, and changes in craving (cigarettes and opioids). These GLM analyses had rsFC as the outcome variable and the following predictor variables: treatment group (iTBS, sham iTBS), time (before iTBS/sham iTBS, after iTBS/sham iTBS), behavioral measure of interest (cigarette cue attentional bias/cigarette craving/ opioid cue attentional bias, or opioid craving), and the treatment group × behavioral measure interactions.

#### Exploratory global rsFC analyses

GLM analyses were conducted with connection-based rsFC in each ROI-ROI pair (105 ROIs in CONN) as dependent variables and independent variables being treatment group, time, and treatment group × time interactions (Detailed in Supplementary Material Section 2).

## Results

### Participants

We screened 36 participants for eligibility. Of these, 23 were enrolled in the study. Four participants withdrew from the study for personal reasons (*n_iTBS_*=2*, n_sham_*=2), and four were lost to follow-up after Day 1 (*n_iTBS_*=2*, n_sham_*=2). Eight participants randomized to iTBS completed all study procedures on Day 2, and five participants randomized to sham iTBS completed all study procedures on Day 2. Two participants randomized to sham iTBS were unable to perform cue attentional bias after the fourth session due to fatigue, but they completed all other study procedures and were therefore included in the analyses.

For rsFC analyses, rsFC data were analyzed from 7 participants in the iTBS group and 7 participants in the sham iTBS group. One participant in the iTBS arm had rsFC data excluded due to poor image quality. Behavioral data were analyzed from eight participants in the iTBS arm and seven participants in the sham iTBS arm. Participants in both arms did not differ in age, sex, FTND scores, or years of opioid use (>**Table 1**>). All participants were on one or more medication(s) for OUD (i.e., methadone, buprenorphine, or naltrexone).

### Seed-based rsFC

There was a significant treatment group × time effect on rsFC between L.dlPFC and a voxel cluster comprising the left anterior supramarginal gyrus (SMG) and the left parietal cortex operculum (*t_12_*=6.37*, beta*=0.40, p<0.001). Planned contrasts showed that rsFC between the L.dlPFC and this SMG cluster increased more after iTBS than after sham iTBS (>**Table 2**>, **Figure 1**). Significant treatment group × time effects were not observed for any rsFC changes from left and right insulae seeds.

**Figure 1.**
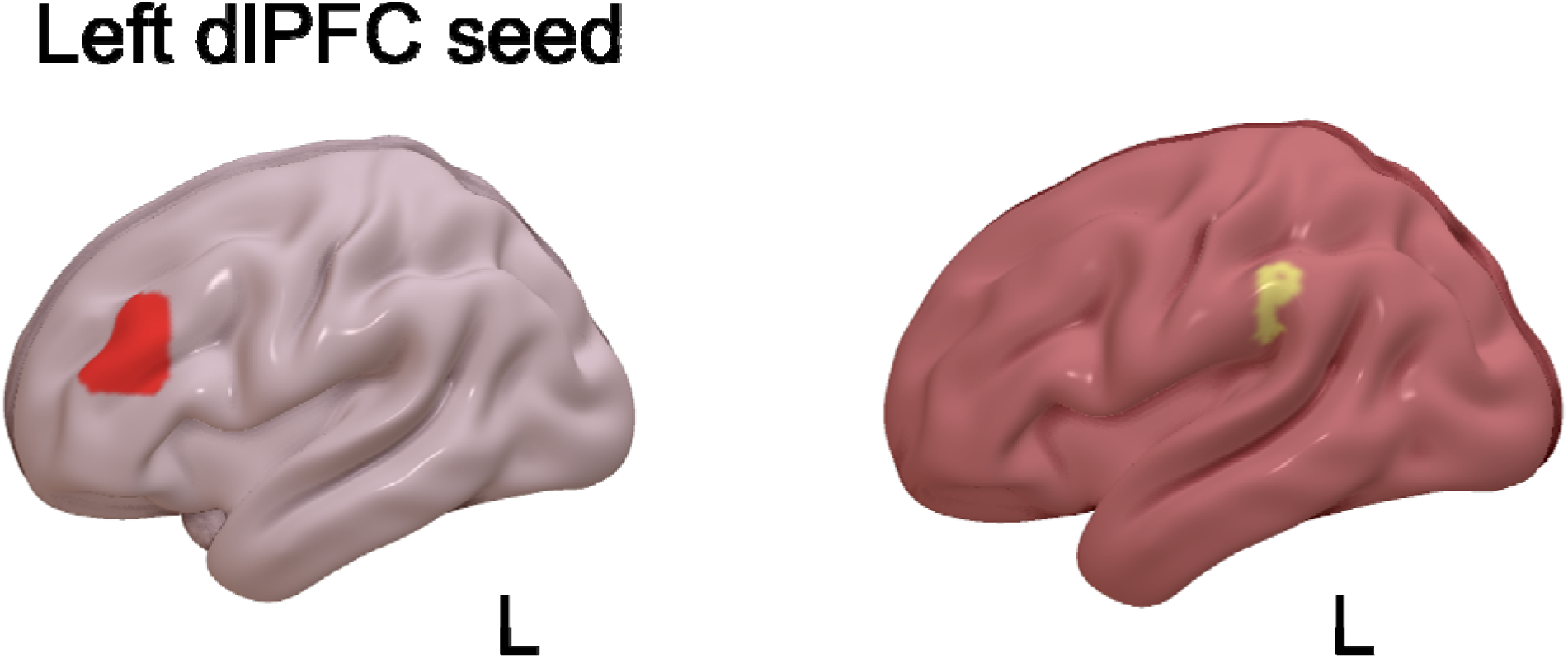
Impact of iTBS/sham iTBS on Seed-Based rsFC (Left dlPFC seed) Figure 1 illustrates increased rsFC between the left dlPFC seed (red) and a voxel cluster (yellow) comprising the left anterior SMG and left parietal operculum, with iTBS compared to sham iTBS.

### Behavioral measures

Cigarette cue attentional bias showed a significant treatment group × session interaction (*t_82_* =2.34*, p*=0.02) (>**Table 3**>, **Figure 2**). Post-hoc comparisons revealed that cigarette cue attentional bias reduced for both cue types (people and paraphernalia) from baseline through sessions one and four in the iTBS arm and increased in the sham iTBS arm (**Figure 2**, **Supplementary Table 4**). Although opioid cue attentional bias showed a nominal reduction in the iTBS arm and an increase in the treatment group receiving the sham iTBS arm, the treatment group*session interaction in an LMEM was not significant (*t_82_*=1.03*, p*=0.31) (**Supplementary Table 1**). Neither cigarette nor opioid craving scores showed significant treatment group × session interactions (*p=0.97 for cigarette craving, p=0.06 for opioid craving*) (**Supplementary Tables 2**> **and 3**>). **Supplementary Table 4** lists the average values for cue attentional bias and craving scores for cigarettes and opioids at three time points in both the iTBS and sham iTBS arms.

**Figure 2.**
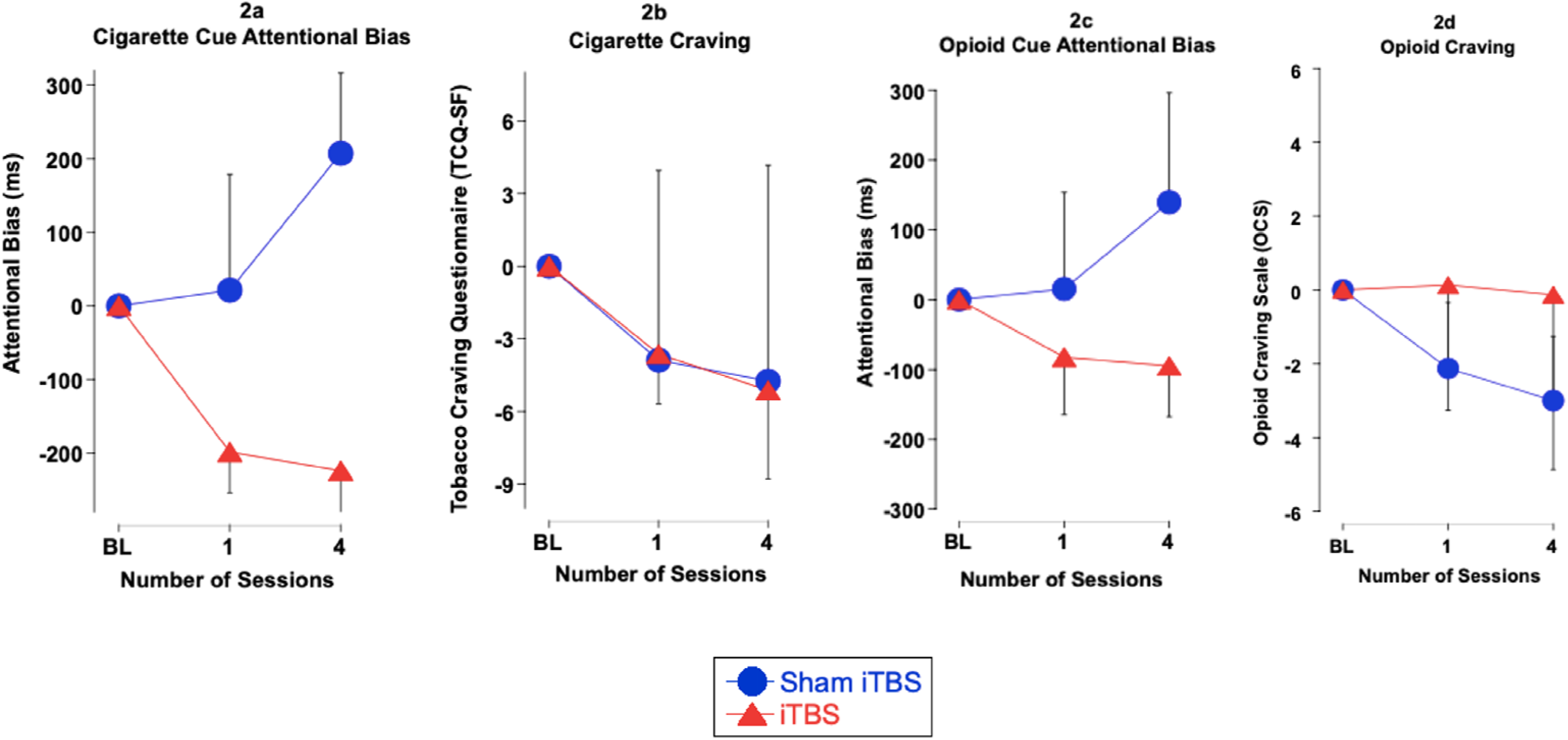
Impact of iTBS/sham iTBS on Cue-Based Attentional Bias and Craving. Figures 2a, 2b, 2c, and 2d illustrate cigarette craving, opioid cue attentional iTBS. The graphs indicate changes fr sessions of iTBS/sham iTBS. Error bars mean (SEM). hanges in cigarette cue attentional bias, bias, and opioid craving with iTBS/sham m baseline (BL), following one and four are indicative of the standard error of the mean (SEM).

### rsFC behavior correlations

GLMs examining the association between rsFC and opioid cue attentional bias, and rsFC and opioid craving, showed significant treatment group × behavioral measure interactions (>**Table 2**>). Compared to sham iTBS, ITBS was associated with reduced positive correlations between opioid cue attentional bias and seed-based rsFC between the left insula and voxel clusters comprising 1) right superior parietal lobule and right postcentral gyrus (*t_11_*=7.25*, beta*=-0.0012, *p-FDR*=0.0002) (iTBS *r_change_*=0.05, sham iTBS *r_change_*=0.11), and 2) right superior frontal gyrus and right precentral gyrus (*t_11_*=9.48*, beta*=-0.0011, *p-FDR*=0.005) (iTBS *r_change_*=0.18, sham iTBS *r_change_*=0.32) (>**Table 2**>, **Figure 3**). Compared to sham iTBS, ITBS increased the positive correlation between opioid cue attentional bias and seed-based rsFC between the left insula and a voxel cluster comprising the left superior division of lateral occipital cortex (*t_11_*=6.75*, beta*=-0.0011, *p-FDR*=0.04) (iTBS *r_change_*=0.10, sham iTBS *r_change_*=0.07) (>**Table 2**>, **Figure 3**).

**Figure 3.**
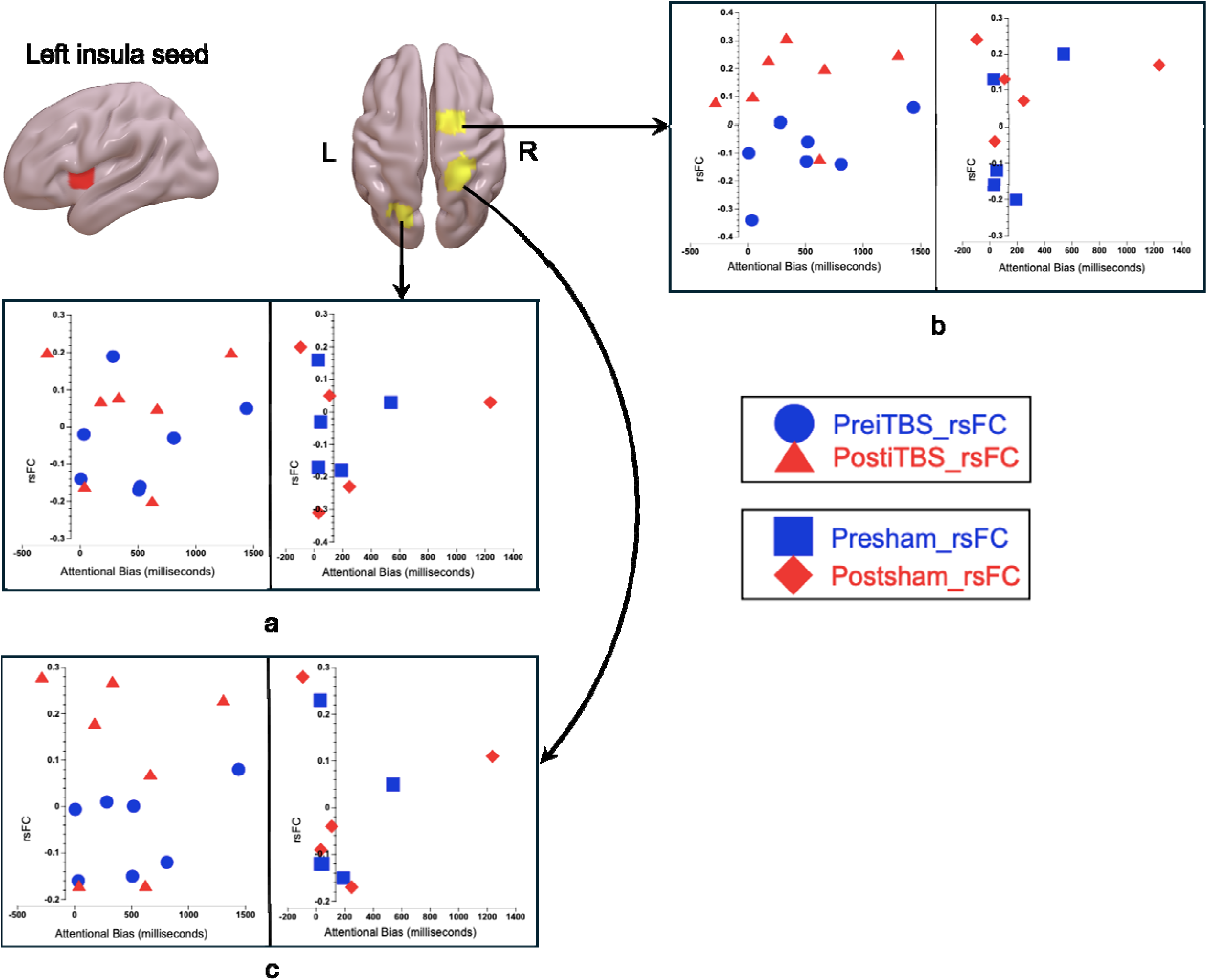
Impact of iTBS/sham iTBS on rsF – Opioid Cue Attentional Bias Correlation. Figure 3 illustrates how iTBS and sham iTBS impacted correlations between seed-based rsFC (left insula seed) and opioid cue attentional bias. a) rsFC between the left insula (red) and a voxel cluster (yellow) comprising the left superior division of lateral occipital cortex demonstrated a higher positive correlation with iTBS (*r_change_* = 0.1) compared to sham iTBS (*r_change_* = 0.07) b) rsFC between the left insula (red) and a voxel cluster (yellow) comprising the right superior frontal gyrus and right precentral gyrus demonstrated a lower positive correlation with iTBS (*r_change_* = 0.05) compared to sham iTBS (*r_change_* = 0.11). c) rsFC between the left insula (red) and a voxel cluster (yellow) comprising the right superior parietal lobule and right postcentral gyrus demonstrated a lower positive correlation with iTBS (*r_change_* = 0.18) compared to sham iTBS (*r_change_* = 0.32).

ITBS demonstrated a negative correlation between opioid craving and seed-based rsFC between the L.dlPFC and a voxel cluster comprising right SMG/middle temporal gyrus compared to sham iTBS (*t_11_*=8.88*, beta*=0.12, *p-FDR*=0.01) (iTBS *r_change_*= -0.01, sham iTBS *r_change_*=0.55) (>**Table 2**>, **Figure 4)**. iTBS demonstrated a positive correlation between opioid craving and seed-based rsFC between the L.dlPFC and a voxel cluster comprising the left lingual gyrus and left intracalcarine cortex compared to sham iTBS, which demonstrated a negative correlation (*t_11_*=5.86*, beta*=0.14, *p* =0.01) (iTBS *r_change_*=0.4, sham iTBS *r_change_*= -0.69) (>**Table 2**>, **Figure 4**). Neither cigarette craving nor cigarette cue attentional bias showed a significant association with seed-based rsFC changes with iTBS/sham iTBS.

**Figure 4.**
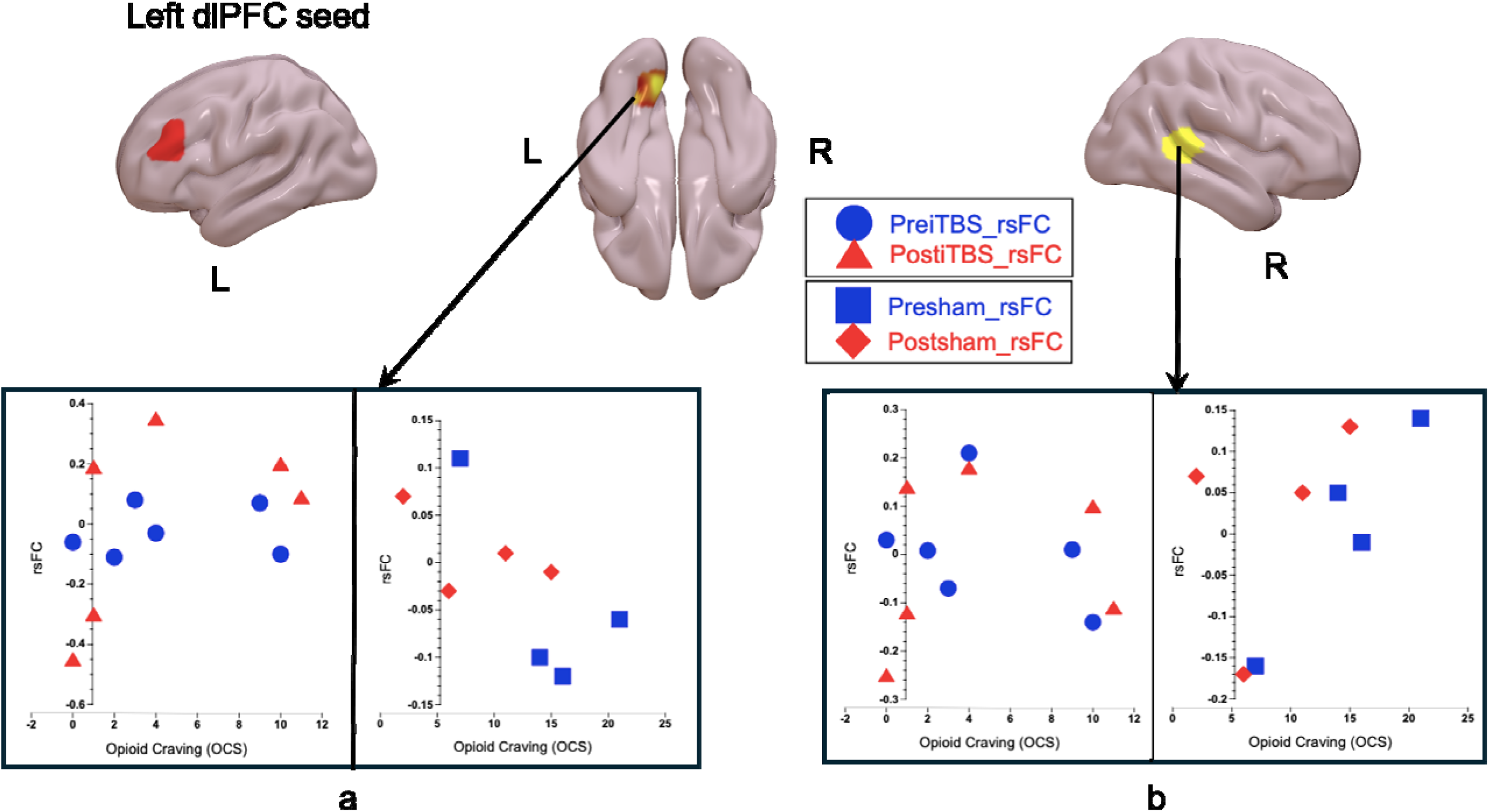
Impact of iTBS/sham iTBS on rsFC – Opioid Craving Correlation. Figure 4 illustrates differences in how iTBS and sham iTBS impacted correlations between changes in seed-based rsFC (left dlPFC seed) and opioid craving. a) rsFC between left dlPFC (red) and a voxel cluster (yellow) comprising the left lingual gyrus/left intracalcarine cortex showed a positive correlation with opioid craving, with iTBS (*r_change_* = 0.4) and a negative correlation with sham iTBS (*r_change_* = -0.69). b) rsFC between the left dlPFC (red) and a temporal gyrus showed a negative correlate 0.01) and a positive correlation with sham iT voxel cluster comprising right SMG/middle n with opioid craving with iTBS (*r_change_* = - S (*r_change_* = 0.55).

### Exploratory Analyses - Global rsFC

Significant treatment group × time effects on ROI-to-ROI rsFC were found in 10 pairs of regions, all of which included SMG (five left SMG and five right SMG [see **Supplemental Table 5**]). Namely, decreases in rsFC between the left and right SMG and cluster in the bilateral striatum (including pallidum) and right thalamus were greater after iTBS than after sham iTBS. The largest treatment group × time effect was observed between the right anterior SMG and the right putamen (*t_12_*=3.75, *beta*=-0.30, *p-FDR*=0.37).

## Discussion

To our knowledge, this is the first pilot study examining the impact of TMS in people with OUD who smoke cigarettes. This is also the first examination of the effects of an accelerated iTBS protocol in people with OUD who smoke cigarettes. Results suggest that four sessions of iTBS targeting the L.dlPFC increased this region’s rsFC with the left SMG, and reduced cigarette cue attentional bias (but not opioid cue bias). Opioid cue attentional bias change correlated with iTBS versus sham iTBS-induced rsFC changes between the left insula and voxel clusters comprising 1) the right superior parietal lobule and right postcentral gyrus, 2) right superior frontal gyrus and right precentral gyrus, and 3) superior division of the left lateral occipital cortex. Opioid craving change correlated with iTBS versus sham iTBS-induced rsFC changes between L.dlPFC and voxel clusters comprising 1) the right SMG/middle temporal gyrus and 2) the left lingual gyrus and left intracalcarine cortex. Although promising, these results should be interpreted cautiously due to the limited sample size.

Of the three seeds, only the L.dlPFC demonstrated a greater change in rsFC with a cluster comprising the left anterior SMG and left parietal operculum after iTBS compared to sham iTBS. This suggests iTBS of the L.dlPFC can modulate the left FPN (comprised of dlPFC and parietal cortices) (Zanto and Gazzaley, 2013). The FPN is involved in working memory, problem-solving, and decision-making (Zanto and Gazzaley, 2013). The FPN has previously demonstrated decreased within-network rsFC in people who smoke compared with those who do not smoke (Weidler et al., 2024, Weiland et al., 2015, Yip et al., 2022). The United Kingdom Biobank dataset showed decreased rsFC between the FPN and two other brain networks, dorsal attention and visual networks, in people who use tobacco products compared to those who do not (Pan et al., 2025). People with tobacco use also showed hypoconnectivity between basal ganglia hubs, in addition to reduced rsFC between basal ganglia hubs and FPN, compared to those without (Pan et al., 2025). In a highly similar vein, our exploratory analyses showed decreased rsFC between the basal ganglia and bilateral SMG (part of the FPN) with iTBS compared to sham iTBS. Overall, the extant literature suggests that rsFC between the FPN and basal ganglia may be crucial for individuals who smoke. Our results indicate that four sessions of accelerated iTBS at the L.dlPFC engaged the left FPN in our study, as well as rsFC between bilateral anterior SMG and basal ganglia.

Earlier studies have shown dose-based engagement of the left FPN by TMS at the L.dlPFC. Interleaved TMS-fMRI in healthy volunteers has demonstrated a dose-related increase in blood oxygen level dependent (BOLD) activity over the L.dlPFC with iTBS (Chang et al., 2024). Six hundred pulses of iTBS delivered over the L.dlPFC at 40% RMT increased BOLD activity over the right dlPFC and not over L.dlPFC. Conversely, the L.dlPFC was engaged with 600 pulses at 80% RMT (Chang et al., 2024). Our earlier study with one session of 1800 pulse iTBS did not engage the left FPN, but four sessions of 1800 pulse iTBS delivered in an accelerated fashion did (Rakesh et al., 2024). Similarly, one iTBS session of 1,800 pulses over the L.dlPFC reduced cigarette craving but not cigarette cue attentional bias compared to sham iTBS (Rakesh et al., 2024). Four sessions of accelerated iTBS had the opposite effect of decreasing cigarette cue attentional bias but did not decrease cigarette craving compared to sham iTBS (Rakesh et al., 2024). The association between attentional bias and craving is weak across substances, as shown by a previous meta-analysis (r=0.2) (Field et al., 2009). The extant literature suggests that both craving and attentional bias have utility in predicting relapse for smoking (Topbas et al., 2025, Vafaie and Kober, 2022). Both facets are associated with differential brain mechanisms, which could have differential effects toward escalating doses of iTBS (Janes et al., 2010, Pan et al., 2025).

Cigarette cue attentional bias and craving did not show significant associations with rsFC. iTBS demonstrated a reduced positive correlation between opioid cue attentional bias and rsFC between the left insula and clusters comprising the right superior frontal gyrus and right superior parietal lobule than sham iTBS. The right superior frontal gyrus overlaps with the right dlPFC. The right dlPFC and right superior parietal lobule comprise the right FPN (Cieslik et al., 2013, Zanto and Gazzaley, 2013). Although opioid cue attentional bias decreased with iTBS compared to sham iTBS, this did not reach statistical significance. Nonetheless, iTBS over the L.dlPFC impacted the relationship between the right FPN - left insula (left salience network node) rsFC, and opioid cue attentional bias. In addition to the changes in the left FPN, the right FPN changes demonstrated by iTBS compared to sham iTBS are essential takeaways from our pilot study.

The insula and FPN are affected in people with opioid use disorder, stabilized on medications for OUD (Ieong and Yuan, 2017, Woisard et al., 2021). Our results demonstrated a negative correlation between opioid craving and seed-based rsFC between the L.dlPFC and right SMG with iTBS, unlike sham iTBS which showed a positive correlation. No previous study has investigated neural correlates of changes in opioid cue attentional bias or opioid craving with iTBS, in people with OUD who smoke. Participants in our study were stabilized on medications for OUD, which could have impacted these neural correlates, in addition to comorbid cigarette smoking. Comorbid OUD and cigarette smoking encompasses a complex interplay of nicotinic cholinergic receptors and opioid receptors (Custodio et al., 2022, Lichenstein et al., 2019). Our pilot study is the first to examine the neural correlates of iTBS in this population. We want to stress the need for more such studies investigating neural correlates to optimize smoking cessation treatments in these patients.

The existing literature suggests that the effects of iTBS depend on the dose and number of sessions, as demonstrated by the differential impact of various iTBS doses on motor-evoked potential amplitude over time (McCalley et al., 2021). Future studies could examine the effect of more days of accelerated iTBS/sham iTBS on attentional bias, craving, and, more importantly, abstinence from smoking.

## Conclusions and Limitations

The small sample size of the present study is a significant limitation. Still, it is comparable to other pilot TMS work that ultimately led to successful outcomes in well-powered clinical trials (Cole et al., 2020, Williams et al., 2018). Future studies should examine the impact of a larger dose of accelerated iTBS at the L.dlPFC (e.g., 20 sessions) on smoking and opioid abstinence and relapse. Continuing to explore the effect of accelerated iTBS in people with OUD who smoke may eventually provide an alternative treatment option for this at-risk population that has repeatedly shown suboptimal response to available treatments.

## Data Availability

All data produced in the present study are available upon reasonable request to the authors

## Acknowledgments

GR is supported by the Junior Research Scholar Award from the Department of Psychiatry, University of Kentucky College of Medicine. TGA is supported by grants R01MH137471 and R61MH132722. SH is supported by grants U54DA058256, R01CA288235, and U01CA275048. RAM is supported by R01MH139648, R01MH111671, and R01MH129832. This pilot study was funded by the Substance Use Priority Research Area (SUPRA) pilot award to GR and the Markey Cancer Center Support Grant (CCSG) Pilot award to SH.

