## Supplementary Material for "Examining the Neural and Behavioral Impact of Accelerated Intermittent Theta Burst Stimulation (iTBS) in People with Opioid Use Disorder (OUD) Who Smoke Tobacco Cigarettes: A Pilot Study"

Section 1. MRI Sequences. Brain MRIs were acquired on a Siemens MAGNETOM 3 Tesla (3T) PRISMA scanner. *Structural MRI*. The structural MRI brain sequence will use a multi-echo magnetization-prepared rapid gradient-echo (MPRAGE) sequence (8.22 minutes) with parameters as follows - FoV 256 mm X 256 mm, slice thickness 0.8 X 0.8 X 0.8 mm^3^, TR 2500 milliseconds (ms), TE – 1.81 ms,3.6 ms, 5.39 ms, 7.18 ms, flip angle 8 degrees(Bookheimer et al., 2019).

*Resting state fMRI*. The resting-state fMRI scan sequence consisted of one run, lasting 6 minutes and 40 seconds, with eyes open and fixated on a crosshair on the scanner screen. We used a multi-slice gradient echo planar imaging (EPI) sequence - FoV 208 mm X 180 mm, slice thickness 2 X 2 X 2 mm^3^, TR 800 ms, TE 37 ms, flip angle 52 degrees (Bookheimer et al., 2019).

Section 2. rsFC analyses. Preprocessing: Functional and anatomical data were preprocessed using a modular preprocessing pipeline (Nieto-Castanon, 2020b), which included realignment with correction of susceptibility distortion interactions, slice timing correction, outlier detection, direct segmentation, MNI-space normalization, and smoothing. Functional data were realigned using SPM realign & unwarp procedure (Andersson et al., 2001), where all scans were coregistered to a reference image (first scan of the first session) using a least squares approach and a six parameter (rigid body) transformation (Karl. J. Friston, 1995), and resampled using b-spline interpolation to correct for motion and magnetic susceptibility interactions. Temporal misalignment between different slices of the functional data (acquired in ascending order) was corrected following SPM slice-timing correction (STC) procedure (R. Henson, 1999, Sladky et al., 2011), using temporal interpolation to resample each slice BOLD timeseries to a common mid-acquisition time. Potential outlier scans were identified using ART (Whitfield-Gabrieli, 2009) as acquisitions with framewise displacement above 0.9 mm or global BOLD signal changes above 5 standard deviations (Nieto-Castanon, 2025, Power et al., 2014). A reference BOLD image was computed for each participant by averaging all scans excluding outliers. Functional and anatomical data were normalized into standard MNI space, segmented into grey matter, white matter, and CSF tissue classes, and resampled to 2 mm isotropic voxels following a direct normalization procedure (Calhoun et al., 2017, Nieto-Castanon, 2025) using SPM unified segmentation and normalization algorithm (Ashburner, 2007, Ashburner and Friston, 2005) with the default IXI-549 tissue probability map template. Last, functional data were smoothed using spatial convolution with a Gaussian kernel of 8 mm full-width half-maximum (FWHM).

Denoising: In addition, functional data were denoised using a standard denoising pipeline (Nieto-Castanon, 2020a) including the regression of potential confounding effects characterized by white matter timeseries (5 CompCor noise components), CSF timeseries (5 CompCor noise components), motion parameters and their first order derivatives (12 factors) (Friston et al., 1996), outlier scans (below 59 factors) (Power et al., 2014), and linear trends (2 factors) within each functional run, followed by bandpass frequency filtering of the BOLD timeseries (Hallquist et al., 2013) between 0.008 Hz and 0.09 Hz. CompCor noise components within white matter and CSF were estimated by computing the average BOLD signal as well as the largest principal components orthogonal to the BOLD average, motion parameters, and outlier scans within each participant's eroded segmentation masks (Behzadi et al., 2007, Chai et al., 2012). From the number of noise terms included in this denoising strategy, the effective degrees of freedom of the BOLD signal after denoising were estimated to range from 275.2 to 303.7 (average 295.6) across all participants (Nieto-Castanon, 2025).

First-level analysis: ROI-to-ROI connectivity matrices were estimated by characterizing the functional connectivity between each pair of regions among 105 Harvard-Oxford atlas ROIs (Nieto-Castanon, 2020c). Functional connectivity strength was represented by Fisher-transformed bivariate correlation coefficients from a general linear model (weighted-GLM) (Nieto-Castanon, 2020d), estimated separately for each pair of ROIs, characterizing the association between their BOLD signal time series. Individual scans were weighted by a boxcar signal characterizing each time point, convolved with an SPM canonical hemodynamic response function, and rectified.

Second-level analyses were performed using a General Linear Model (GLM) (Worsley et al., 1996). For each connection, a separate GLM was estimated, with first-level connectivity measures at that connection as dependent variables and the treatment group, time, and group-time interactions as independent variables. Connection-level hypotheses were evaluated using multivariate parametric statistics with random effects across participants and sample covariance estimation across multiple measurements. Inferences were performed at the level of individual clusters (groups of similar connections). Cluster-level inferences were based on parametric statistics within and between each pair of ROIs (Jafri et al., 2008), considering ROI-to-ROI anatomical proximity and functional similarity (Nieto-Castanon, 2020d). Results were thresholded using a combination of a p < 0.001 voxel-level threshold and a familywise p-value corrected with (false discovery rate) FDR < 0.05 cluster-level threshold.

Table 1. LMEM. Opioid cue attentional bias as the DV

Opioid cue attentional bias~1+ treatment group (iTBS / sham iTBS) + session (one versus four) + treatment group*session + (1|ParticipantID) (R^2^ = 0.73)

Opioid cue attentional bias is the DV, and the following are predictor variables: treatment group (iTBS / sham iTBS), session (baseline, after one session, and after four sessions), and participant identification number (Participant ID).

| Name | Estimate (Beta) | SE | T Statistic | Degrees of freedom | p value |
| --- | --- | --- | --- | --- | --- |
| Intercept | 188.76 | 165.84 | 1.14 | 82.00 | 0.26 |
| Group | 259.2 | 226.95 | 1.14 | 82.00 | 0.26 |
| Session | 26.13 | 54.92 | 0.48 | 82.00 | 0.64 |
| Group × session | -73.61 | 71.43 | -1.03 | 82.00 | 0.31 |

Table 2. LMEM. Cigarette craving as the DV

Cigarette craving (TCQ-SF)~1+ treatment group (iTBS / sham iTBS) + session+ treatment group*session + (1|ParticipantID) (R^2^ = 0.43)

Cigarette craving is the DV, and the following are predictor variables: treatment group (iTBS / sham iTBS), session (baseline, after one session, and after four sessions), and participant identification number (Participant ID).

| Name | Estimate (Beta) | SE | T Statistic | Degrees of freedom | p value |
| --- | --- | --- | --- | --- | --- |
| Intercept | 52.57 | 4.94 | 10.65 | 41.00 | <0.005 |
| Group | 4.82 | 6.76 | 0.71 | 41.00 | 0.48 |
| Session | -2.71 | 2.73 | -0.99 | 41.00 | 0.33 |
| Group × session | 0.15 | 3.74 | 0.04 | 41.00 | 0.97 |

Table 3. LMEM. Opioid craving as the DV

Opioid craving (OCS)~1+ treatment group (iTBS / sham iTBS) + session + treatment group*session + (1|ParticipantID) (R^2^ = 0.87)

Opioid craving is the DV, and the following are predictor variables: treatment group (iTBS / sham iTBS), session (baseline, after one session, and after four sessions), and participant identification number (Participant ID).

| Name | Estimate (Beta) | SE | T Statistic | Degrees of freedom | p value |
| --- | --- | --- | --- | --- | --- |
| Intercept | 8.05 | 2.08 | 3.86 | 41.00 | 0.0003 |
| Group | -4.49 | 2.85 | -1.57 | 41.00 | 0.12 |
| Session | -1.71 | 0.61 | -2.79 | 41.00 | 0.01 |
| Group × session | 1.65 | 0.84 | 1.96 | 41.00 | 0.06 |

*- Significant

Table 4 – Planned Comparisons of Attentional Bias and Craving for Cigarettes/Opioids Across Groups and Time Points

| Treatment Arm | Baseline | After 1 Session | After 4 sessions |
| --- | --- | --- | --- |
| Cigarette Cue Attentional Bias | | | |
| iTBS | 534.01 (104.45) | 335.3 (105.74) | 310.04 (74.9) |
| Sham iTBS | 298.97 (76.55) | 320.38 (157.7) | 534.15 (122.33) |
| Opioid Cue Attentional Bias | | | |
| iTBS | 459.56 (124.18) | 377.31 (137.45) | 364.60 (128.76) |
| Sham iTBS | 192.37 (65.15) | 207.68 (151.97) | 306.27 (168.51) |
| Cigarette Craving (TCQ-SF) | | | |
| iTBS | 57.8 (5.3) | 54.1 (6.3) | 52.6 (6.3) |
| Sham iTBS | 53.1 (6.1) | 48.7 (4.9) | 47.7 (5.1) |
| Opioid craving (OCS) | | | |
| iTBS | 3.5 (1.6) | 3.6 (1.6) | 3.4 (1.6) |
| Sham iTBS | 8.3 (3.3) | 5.9 (2.8) | 4.9 (2.3) |

Table 5: Global rsFC analyses: Regions with Significant rsFC Differences Across Groups and Time Points

| **Region** | **Beta** | **T-statistic** |
| --- | --- | --- |
| Post minus Pre, iTBS minus sham iTBS | | |
| Right putamen-right anterior SMG | -0.3 | 3.75 |
| Right thalamus-right anterior SMG | -0.28 | 4.32 |
| Left caudate-left anterior SMG | -0.28 | 3.33 |
| Right caudate-left anterior SMG | -0.26 | 2.58 |
| Left caudate-right anterior SMG | -0.24 | 2.23 |
| Left putamen-left anterior SMG | -0.22 | 3.36 |
| Right thalamus-left anterior SMG | -0.21 | 2.46 |
| Right putamen- left anterior SMG | -0.2 | 2.8 |
| Right pallidum-right anterior SMG | -0.18 | 2.38 |
| Left putamen-right anterior SMG | -0.15 | 2.21 |
